# A double-blind, sham-controlled, trial of home-administered rhythmic 10Hz median nerve stimulation for the reduction of tics, and suppression of the urge-to-tic, in individuals with Tourette syndrome and chronic tic disorder

**DOI:** 10.1101/2023.03.06.23286799

**Authors:** Barbara Morera Maiquez, Caitlin Smith, Katherine Dyke, Chia-Ping Chou, Belinda Kasbia, Ciara McCready, Hannah Wright, Jessica K. Jackson, Isabel Farr, Erika Badinger, Georgina M. Jackson, Stephen R. Jackson

## Abstract

Tourette syndrome (TS) and chronic tic disorder (CTD) are neurological disorders of childhood onset characterised by the occurrence of tics; repetitive, purposeless, movements or vocalisations of short duration which can occur many times throughout a day. Currently, effective treatment for tic disorders is an area of considerable unmet clinical need. We aimed to evaluate the efficacy of a home-administered neuromodulation treatment for tics involving the delivery of rhythmic pulse trains of median nerve stimulation (MNS) delivered via a wearable ‘watch-like’ device worn at the wrist. We conducted a UK-wide parallel double-blind sham-controlled trial for the reduction of tics in individuals with tic disorder. The device was programmed to deliver rhythmic (10Hz) trains of low-intensity (1-19 mA) electrical stimulation to the median nerve for a pre-determined duration each day, and was intended to be used by each participant in their home once each day, 5 days each week, for a period of 4 weeks. Between 18^th^ March 2022 and 26^th^ September 2022 135 participants (45 per group) were initially allocated, using stratified randomisation, to one of the following groups; active stimulation; sham stimulation; or a to waitlist (i.e., treatment as usual) control group. Recruited participants were individuals with confirmed or suspected TS/CTD aged 12 years of age or upward with moderate to severe tics. Researchers involved in the collection or processing of measurement outcomes and assessing the outcomes, as well as participants in the active and sham groups and their legal guardians were all blind to the group allocation. The primary outcome measure used to assess the ‘offline’ or treatment effect of stimulation was the Yale Global Tic Severity Scale - Total Tic Severity Score (YGTSS-TTSS) assessed at the conclusion of 4-weeks of stimulation. The primary outcome measure used to assess the ‘online’ effects of stimulation was tic frequency, measured as the number of tics per minute (TPM) observed, based upon blind analysis of daily video recordings obtained while stimulation was delivered. The results demonstrated that after 4-weeks stimulation, tic severity (YGTSS-TTSS) had reduced by 7.1 points (35% reduction) for the active stimulation group compared to 2.13/2.11points for the sham stimulation and waitlist control groups. The reduction in YGTSS-TTSS for the active stimulation group was substantially larger, clinically meaningful (effect size = 0.5), and statistically significant (p = 0.02) compared to both the sham stimulation and waitlist control groups, which did not differ from one another (effect-size = -0.03). Furthermore, blind analyses of video recordings demonstrated that tic frequency (tics per minute) reduced substantially (−15.6 TPM) during active stimulation compared to sham stimulation (−7.7 TPM). This difference represents a statistically significant (p < 0.03) and clinically meaningful reduction in tic frequency (> 25% reduction: effect-size = 0.3). These findings indicate that home-administered rhythmic MNS delivered through a wearable wrist-worn device has potential as an effective community-based treatment for tic disorders.

## Introduction

Tourette syndrome (TS) and chronic tic disorder (CTD) are neurodevelopmental disorders that are found in the majority of cultures worldwide (Robertson, Eapen, & Cavanna, 2009) and impact approximately 1% of 5-18 year olds (Cohen, Leckman, & Bloch, 2013). Both TS and CTD are characterised by the presence of tics, which are repetitive, purposeless, movements or vocalisations of short duration which can occur many times throughout a day. Tics are highly varied and can range from simple movements and/or vocalisations such as mild eye blinking and throat clearing; to more complex sequences of movement and behaviour, including, mimicking sounds or blurting out obscenities. The majority of adults and adolescents with TS/CTD also experience premonitory urges (PU). PU are uncomfortable sensory phenomena, often described as feelings of discomfort or pressure which can be temporally reduced after tic execution (Cohen et al., 2013). Many individuals with TS/CTD will also experience one or more co-occurring conditions, with the most common being attention deficit hyperactivity disorder (ADHD) and obsessive compulsive disorder (OCD) (Freeman et al., 2000).

Tics can have a substantially negative impact on an individual’s day-to-day life, with social, occupational/academic, and psychological well-being affected (Conelea et al., 2011; Conelea et al., 2013). Rates of depression are higher in people with TS/CTD than the general population (Robertson, 2006), as is the risk of dying by, or attempting suicide (de la Cruz et al., 2017). Despite these concerning statistics, access to support and treatments for TS/CTD is often limited and sub-optimal.

The two main evidence-based approaches to treating tics are behavioural therapies and medication (Roessner, 2011; Whittington et al., 2016). A recent systematic review found that over half of young people with TS had received medication to help with their tics; however, for many the associated side effects (which can include sedation and weight gain) outweigh the potential benefits (see Hollis et al. (2016) for review). Behavioural treatments such as habit reversal therapy (Azrin & Nunn, 1973) and extensions of this, such as comprehensive behavioural intervention for tics (Piacentini et al., 2010), have been shown to be effective treatments. However, access to specialists who are able to provide this type of therapy is often limited, for example, a UK based study found that approximately 25% of young people with TS had access to behavioural interventions, despite 76% of parents indicating that they would like access to this treatment for their child (Cuenca et al., 2015).

Dysfunction within cortical – striatal – thalamic – cortical (CSTC) circuits has been heavily implicated in the pathophysiology of tic disorders (Greene, Schlaggar, & Black, 2015; Mink, 2006). In particular, it is thought that the dysfunction in CSTC leads to spontaneous firing of the striatum, which releases the thalamus from tonic inhibition, resulting in the increased excitability of the sensorimotor cortex leading to the generation of tics (Worbe et al., 2013; Xu et al., 2015).

In response to the demand for non-pharmacological treatments, numerous studies have utilised non-invasive brain stimulation (NIBS) approaches, with the aim of readdressing imbalances within cortical excitability as a means to reduce tics. The majority of this work has focused on two techniques: repetitive transcranial magnetic stimulation (rTMS) and transcranial direct current stimulation (tDCS), and targeted primary (M1) and supplementary (SMA) motor regions. Both tDCS and rTMS have been shown to modulate cortical excitability during, and shortly after stimulation, via processes thought to be similar to long term depression (LTD) and long term potentiation (LTP) (Huang, Chen, Rothwell, & Wen, 2007; Nitsche et al., 2003). While some studies have shown significant tic reductions following several sessions of rTMS (Hsu, Wang, & Lin, 2018), the overall picture of results remains somewhat varied, and large scale sham-controlled trials are needed. Furthermore, access to rTMS treatment is typically restricted to research studies, which often involve up to two weeks of consecutive sessions conducted within a clinic/research facility, hence, this approach puts heavy demands on time and resources for both participants and researchers. tDCS is a more portable form of NIBS, and has been trialled in several conditions for home use application (Charvet, Shaw, Bikson, Woods, & Knotkova, 2020); however, relatively few studies of the effectiveness of tDCS on tics have been conducted, and as with rTMS there remains a need for larger scale studies with sufficient control parameters incorporated into the design (Fregni et al., 2020).

A limitation of both tDCS and TMS, is that these approaches both require application of stimulation to the scalp in order to reach the cortical targets below. For optimal targeting of specified brain regions an MRI may be necessary for current flow modelling and optimising coil/electrode placement, this is neither cheap nor easily acquired in groups with movement disorders. Furthermore, the nature of transcranial stimulation is that it is not discrete, and it is unlikely that this technology could be developed into an approach that individuals could use within the home.

An alternative to transcranial stimulation, is to stimulate the peripheral nervous system, which can lead to targeted responses within cortical regions. In recent work using electroencephalography (EEG) we have successfully shown that pulses of electrical stimulation delivered to the median nerve are capable of entraining neural oscillations within the sensorimotor cortex (Morera Maiquez et al., 2020). Houlgreave and colleagues have shown the same effect in healthy adults using magnetoencephalography (MEG) (Houlgreave et al, 2020). Specifically, they demonstrated entrainment of oscillatory activity within the (8-14Hz) alpha/mu band and the (15-30Hz) beta band, which are associated with sensorimotor function (Armstrong, Sale, & Cunnington, 2018). Importantly, it has been shown that when median nerve stimulation (MNS) was administered to people with TS/CTD, their urge-to-tic and the occurrence of their tics substantially reduced (Morera Maiquez et al., 2020). The Morera Maiquez et al. study was conducted with 19 individuals and assessed the immediate impact of MNS applied under experimental conditions. We now build on the encouraging results of this work, by conducting a UK-wide parallel double-blind study in which the potential beneficial effects of numerous sessions of MNS, delivered in the home environment, were evaluated.

### Our key aims of the study were as follows

1. Evaluate the ‘online’ effects of MNS by exploring alterations in *tic frequency* during MNS, through the use of video recordings collected during the delivery of stimulation.

2. Evaluate the ‘offline’ or treatment effects of MNS tic severity.

The primary hypotheses are: (a) that active rhythmic MNS will lead to a reduction in tic severity compared to sham stimulation and a waitlist (treatment-as-usual) control group; (b) that active rhythmic MNS will lead to a reduction in tic frequency compared to both sham stimulation; and (c) that rhythmic MNS can be successfully delivered using a wearable ‘watch-like’ device in a home setting.

## Methods

### Setting

This trial assessed the effectiveness of 4-weeks of daily, home administered, rhythmic mu-band MNS as a treatment for tic disorders. Following an initial baseline assessment/training visit at the University of Nottingham, the trial was conducted within each participant’s home, with remote online supervision.

### Recruitment

We targeted an enrolment of 135 participants in the trial. Participants were recruited from our existing database of volunteers and through the UK charity *Tourettes Action*.

### Inclusion criteria

1. Ages 12 years upward.

2. Confirmed or suspected Tourette syndrome/Chronic tic disorder. With moderate-severe tics, indicated by a total tic score > 15 on the Yale global tic severity scale (YGTSS), or total tic score > 10 if only motor/ vocal tics present.

3. No change in medication for tics or tic-related treatment in the last 2 months. Participants to confirm this during telephone screening.

4. Broadband internet access & electronic device for completion of online materials. For a subset of participants, a device with a camera will also be required.

5. Ability to travel to the University of Nottingham for one onsite visit.

6. Participant is willing and able to give informed consent for participation in the clinical investigation.

7. Able (in the Investigator’s opinion) and willing to comply with all clinical investigation requirements.

8. Resident in the UK.

### Exclusion criteria

1. Current diagnosis of epilepsy.

2. Participant or participants guardian (if under 16) unable to read/write in English.

3. Participants will be excluded from the trial if they find the stimulation too uncomfortable during the in-person baseline assessment visit.

4. Individuals with implanted electronic devices (e.g. pacemakers, insulin pump, implantable cardioverter defibrillator, neurostimulators)

5. Individuals sharing the household with an individual with implanted electronic devices (e.g. pacemakers, insulin pump, implantable cardioverter defibrillator, neurostimulators)

6. Individuals with a current/recent diagnosis or symptoms of SARS-CoV-2 were not be invited to visit the university until it was safe for them to do so (2 weeks following positive test).

7. Diagnosis of non-verbal autism or similar condition which would affect ability to give informed consent to take part in the study.

8. Pregnant women.

9. Participants who have participated in previous research studies involving median nerve stimulation

10. Participants aged over 90 years old

#### Initial Screening

Individuals who had indicated an interest in taking part in the study were contacted by a member of the research team to arrange a telephone screening interview. Trial eligibility was then established during this interview using the inclusion & exclusion criteria outlined above. Suitable participants were informed about each step of the trial and the randomisation procedure; they were provided the opportunity to ask questions about the study and received a detailed information sheet. Written informed consent was taken prior to the initial screening videocall and again prior to enrolment in the full trial, if the participant was found to be eligible, using an online form.

### Randomization and blinding

Participants were randomly allocated into three groups (ratio: 1:1:1): active rhythmic median nerve stimulation (rMNS) stimulation, sham rMNS stimulation, and a waitlist (i.e., treatment-as-usual and no stimulation). In order to minimize any differences in age, sex and tic severity between groups, a stratified randomisation procedure was followed in which individuals allocated to each group were matched for age, sex and tic severity. The devices used to deliver active and sham stimulation were exactly the same to ensure that researchers were blind to a participant’s group allocation. The first 20 participants who were randomly assigned to each group, and who exhibited very frequent tics (i.e. tic-free intervals that were typically no longer than 5min), were selected as a subgroup who were required to additionally provide video recordings immediately prior to, during, and immediately after they received stimulation. These recordings were used to assess online effects of rMNS stimulation.

Importantly, the member of the research team allocating participants to each condition was not involved in either the collection or processing of measurement outcomes (questionnaire/ video data). Similarly, the researchers responsible for programming the wearable MNS devices, to deliver either sham/active stimulation, were not involved in the collection or processing of measurement outcomes (questionnaire/video data). All other members of the research team, participants, and legal guardians were entirely blind to sham/active group allocation. Participants allocated to the waitlist group were not blind to the type of stimulation they would go on to receive at the conclusion of the trial (i.e., all participants initially allocated to the waitlist group went on to receive active rMNS at the conclusion of the trial).

### Baseline data collection and visit

Prior to any further measures being collected, a subset of 16 participants from each group were asked to video record themselves during restful activity (such as watching television) for 5 minutes on 5 consecutive days. The purpose of this was to obtain a baseline of tics for these individuals prior to any intervention. Participants were instructed how to video record themselves, and how share the video with the research team, through a videocall training session.

All participants allocated to the active and sham groups were then invited to the University of Nottingham. During this visit, the participants received their stimulation device and were trained on its correct placement and use. In order to ensure participant’s comfort with the stimulation, a practice session was performed. If the participant experienced significant discomfort, they were withdrawn from the trial. On the same day, demographic information along with primary and secondary measures were collected using various questionnaires and structured interviews. Participants in the waitlist group also completed these measurements online and through a video call.

Participants in the active and sham stimulation groups returned home with the device and were instructed to commence stimulation sessions on a Monday within 3 weeks of their visit.

### Trial design

A randomised, parallel, double-blind, sham-controlled design was used for this trial (active stimulation vs. sham stimulation condition). The trial also included an open label, waitlist (treatment-as-usual) control condition, in which participants experienced treatment as usual prior to receiving active stimulation at the conclusion of the trial. After screening and attending a baseline visit at the University of Nottingham, the active and sham groups were asked to use the stimulation daily from Monday to Friday, within their own homes for 4 consecutive weeks. The waitlist group did not receive stimulation for the first four weeks, but were then provided with devices set for active stimulation for home administration (i.e., similar to that provided for the active stimulation group). However, unlike the active stimulation group, the participants in the waitlist group were not blind to the stimulation type they would receive. Outcome measurements based upon questionnaires and semi-structured interviews were obtained at baseline, and weekly during the 4 weeks of stimulation, and then at 3-month and 6-month follow-up (see schematic in Figure 1). Parents of participants under 18 years old were asked to be present during the online and video call measures, and while the participant used the stimulation device.

**Figure 1:**
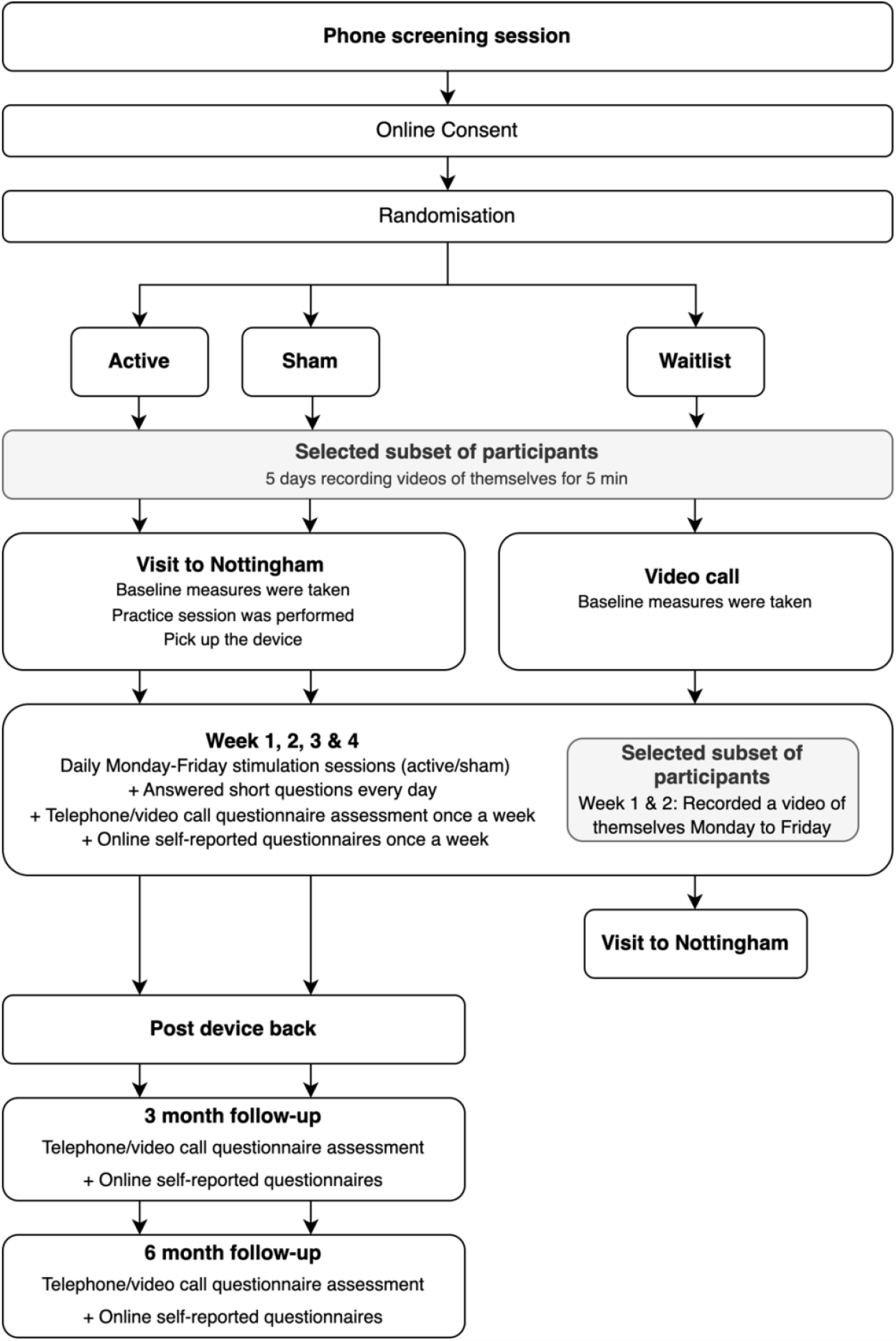
schematic flow diagram illustrating trial design.

As noted previously, a subset of 16 participants from each group were asked to record short video recordings of their tics. Initially this involved recording for 5 minutes on 5 consecutive days to create a baseline measure of tic severity. Then, during the first two weeks of stimulation use, the subset of participants in the active and sham stimulation groups were asked to video record themselves immediately before stimulation (5 mins), during stimulation (14 mins) and immediately after stimulation (5 mins). The subset of 16 participants on the waitlist group were asked to record 5 minute videos of themselves over the same time period. Video recording took place on weekdays (Monday-Friday) at approximately the same time of day. Participants were requested to collect these video recordings while sitting at a table and engaged in a passive, restful, activity such as watching television.

After 4 weeks of device use, participants were asked to return the device. Questionnaires and semi-structured interviews to assess primary and secondary outcome measures were repeated at 3 and 6 months after the start of the stimulation sessions. At the end of the trial, participants were fully debriefed and informed into which group they had been allocated. Here we are reporting data collected at baseline and during the 4-week stimulation phase of the trial. Data for the 3-month and 6-month follow-up will be reported separately at a later point in a further publication.

A schematic flow diagram of the trial design is shown in *Figure 1*.

### Delivery and monitoring of MNS

We used wearable stimulation devices specially designed and built for this trial, with the aim of delivering rMNS stimulation in a similar manner to that reported in a previous study (Morera et al., 2020). These devices were lightweight, wireless and easy to operate. Importantly, participants were not able to edit the device parameters once these had been programmed, thus the duration, frequency and intensity of stimulation remained fixed throughout the trial for each participant. To ensure that all participants underwent daily sessions of stimulation, the device incorporated software that updated the research team after each use. The device was also restricted so that it could only be used once each day. To ensure that the participants were wearing the device during stimulation, the device only operated if it was correctly attached to the wrist.

The intensity of stimulation (1-19 mA) was individualised for each participant based on the approach previously used in Morera et al. (2020). Specifically, the stimulation threshold for each participant was determined by delivering single pulses to the wrist at an increasing intensity until a visible contraction in the thenar muscle was observed. In the active group, a session of stimulation consisted of delivering rhythmic pulse trains of MNS at a frequency of 10Hz in which each pulse was of 200 µs (i.e., 0.2 ms) duration and was delivered at 120% of motor threshold, in bursts of 2 minutes of stimulation followed by 1 minute of no stimulation. This was repeated 5 times, lasting 14 minutes in total. Stimulation was delivered on the wrist of their right hand. In the sham group, the same pattern and total duration of stimulation was applied; however, for the first 15 seconds of each session only, stimulation was delivered at 120% motor threshold, after which it was ramped down to 50% of motor threshold thereafter. This approach to sham stimulation ensured participants initially felt the stimulation prior to it being reduced. Pilot data using magnetoencephalography (MEG) demonstrated that MNS delivered at 50% of stimulation threshold does not cause entrainment of neural oscillations.

### Sample size

Based on previous studies (e.g. Debes et al. (2015); Jankovic et al. (2016); Morera Maiquez et al. (2020); Stenner, Baumgaertel, Heinze, Ganos, and Muller-Vahl (2018)) we estimated that a 6-point reduction (i.e., 25% reduction) in the YGTSS total tic severity score (YGTSS-TTSS) would indicate a clinically meaningful improvement in tic severity (Jeon et al., 2013). Since we predicted that there would be a clinical improvement in the active group compared to the sham and waitlist groups, a one-sided type I error of 2.5% with a 90% power required a total sample of 39 participants per group. Furthermore, in order to allow for a 13% dropout, 45 participants per group, and a total of 135 participants, were recruited for the trial. The sample size for the study exploring the online effects of the stimulation using video data was established as 16 participants per group. This was treated as exploratory analyses.

### Measures

A combination of self-report questionnaires, semi-structured interviews, and video recording of tic frequency were used. These were collected during the baseline in-person visit, through online forms or through video/telephone calls. All interview-based measures were completed by trained researchers. Where possible, the same researcher assessed the same participant throughout the trial. Video/telephone calls were recorded for quality checks. Any changes in tic medication/treatment during the trial were noted. A schematic flow diagram of the trial measures is shown in Supplementary Figure 1.

## Demographic measures

To assess characteristics of the participant sample, the following measures were taken during the baseline assessment. These measures were collected in person for the sham and active groups during their visit to the University of Nottingham. Measures were collected using video call and through online forms for those in the waitlist group, with the exception of the IQ measure which was collected in-person when participants visited the university to start the open label active phase of the study.

- The Autism-spectrum quotient [AQ]/autism-spectrum quotient adolescent (Baron-Cohen, Hoekstra, Knickmeyer, & Wheelwright, 2006; Baron-Cohen, Wheelwright, Skinner, Martin, & Clubley, 2001): 50-item self-report measure giving an indication of autistic traits.
- Edinburgh Handedness Inventory–short form [EHI] (Veale et al, 2014): 4-item self-report measure to assess hand dominance.
- Wechsler’s abbreviated scale of intelligence, two subtests form (WASI-II). (Wechsler, 2011): researcher lead assessment of matrix reasoning and vocabulary used to provide a rough IQ estimate.
- Becks depression inventory (Beck, Erbaugh, Ward, Mock, & Mendelsohn, 1961): 21-item self-report questionnaire to assess symptoms of depression.
- Age appropriate measures were used to assess symptoms of ADHD: the World health organisation adults ADHD self-report scale (ASRS) (Kessler et al., 2005) or Conners comprehensive behavioural ratings scale self-report (Conners, 2008); dependant on the age of participant.
- Estimated age of tic onset.
- Any previous treatments received to help with tics.
- Any confirmed current diagnoses of co-occurring neuropsychiatric conditions.
- Current prescribed medications for tic disorders and/or co-occurring neuropsychiatric conditions.

## Primary outcome measures

The primary outcome measure was the Yale Global Tic Severity Scale revised [YGTSS-R] total tic severity score [TTSS] (McGuire et al. 2018). The YGTSS is a validated, researcher-administered, semi-structured symptom checklist of 46 tic disorder symptoms occurring within the last week. The YGTSS-TTSS includes subscales for tic number, frequency, intensity, complexity and interference. These subscales can be combined to form total motor tic score and total phonic tic score, each with a possible rating of 0-25. By combining the two scores the total tic severity score is calculated. The YGTSS was administered by a trained researcher blind to the experimental group of each participant. The first YGTSS measure was conducted in person (sham/active stimulation groups) or through video call (open label waitlist group). Subsequent measures of the YGTSS were completed by video call at weeks 1-4, at 3 months follow-up and 6 months follow-up.

## Secondary outcome measures

Secondary outcome measures were taken at various time points, including at baseline, weeks 1-4, and at 3 and 6 month follow-up.

- Premonitory Urge for Tics Scale-Revised [PUTS-R] (Baumung et al., 2020): The PUTS-R is a 24 item self-report instrument which is specifically designed to measure the current frequency of different types of premonitory urges in patients with tic disorders. We used the total score on PUTS-R as a primary outcome measure to assess changes in premonitory urge.
- (Children’s) Yale-Brown Obsessive-Compulsive Scale (C)Y-BOCS (Goodman et al., 1989; Scahill et al., 1997): The age appropriate version of this semi-structured interview was used to assess symptoms of OCD. The first part of the scale involves assessing what potential obsessions/compulsions an individual has experienced over the course of the past week, followed by assessment of the time spent, interference and distress caused by, ability to resist, and control over obsessions/compulsions.
- Gilles de la Tourette Syndrome – Quality of Life scale (TS-QoL) (Cavanna et al., 2013; Cavanna et al., 2008): The age appropriate version of this 27-item semi-structured interview consisting of four subscales (psychological, physical, obsessive-compulsive and cognitive) which can be combined to give a single measure indicating overall quality of life was used. The questionnaire also includes a measure of current satisfaction with life using a visual analogue scale (VAS).
- Becks anxiety scale (BAS) (Beck, Epstein, Brown, & Steer, 1988) 21-item self-report questionnaire to assess symptoms of anxiety.
- Yale Global Tic Severity Scale revised [YGTSS-R] impairment score (McGuire et al. 2018). The impairment score is a value of 0-50 given by participants in response to a question about the level of distress and impairment they feel as a result of their tics in areas of daily life including interpersonal, academic and occupational.

Note, data from all measures collected at 3-month and 6-month follow-up will be reported separately at a later time.

## Video recording of tics

A subset of participants were asked to record videos of themselves, and share these daily with the research team. The videos included the participant’s face and upper body, but did not include the hand being stimulated at rest. Each video was reviewed daily for quality by a member of the research team who was not involved in either the collection or processing of measurement outcomes (questionnaire/ video data), to ensure it met the following criteria:

∘ The room is well-lit and the participant’s face and facial tics can be clearly seen.
∘ The participant is sitting at a table with their recording device in front of them.
∘ The participant’s face and upper body are in the video.
∘ The participant’s hands are not in the video.
∘ The participant’s whole face is visible throughout the entire recording.
∘ The participant is quiet and not speaking or singing during the recording.
∘ The room is in silence during the recording, making vocal tics easy to hear.
∘ The participant is relaxed and is not engaging in activities involving voluntary movements, such as such as using their phone.
∘ The participant has recorded approximately 5 minutes before starting the device and continues to record for approximately 5 minutes after the end of the stimulation.

### Statistical analyses and data processing

Our preference is to report effect sizes and to use non-parametric permutation statistics and/or Bayesian hypothesis testing where appropriate to circumvent issues associated with changes in sample size (i.e., drop-out or missing data) or sample variance (to be expected if the intervention is effective for some but not all participants). Statistical analysis using parametric analyses (e.g., ANOVA) are also used where this is appropriate.

### Demographic information

Demographic data are summarized by group. Continuous data within groups is reported as mean and standard deviation (SD), median and inter-quartile range (IQR). Categorical data within groups is reported as a percentage.

### Primary outcome analyses: Change in tics indicated by YGTSS-TTTS, YGTSS-motor and YGTSS-phonic scores

To assess the effect of 4 weeks of daily rMNS treatment on tic severity, we calculated individual difference scores for each measurement by subtracting each individual’s YGTSS score measured at baseline from their score measured at the end of the 4-week stimulation period (i.e. week 4). Then, to assess between-group differences in reduction in tic severity, we conducted planned contrasts (i.e., active vs. sham; active vs. waitlist, and sham vs. waitlist) of the mean difference measurements using non-parametric permutation testing.

### Secondary outcome analyses

Individual difference scores were computed (as above) for secondary outcome measures, specifically: YGTSS-impairment score; premonitory urge (PUTS-R) score; and OCD symptoms (CY-BOCS). Between-group differences are assessed as outlined above.

## Exploratory analyses

Multiple regression analyses were used to explore potential predictive relationships between change in tic severity (measured at week 4) and measures of co-occurring conditions/symptoms measured at baseline. Specifically, the variables: group membership, OCD total score (CY-BOCS-total), OCD obsessions score (CY-BOCS-obsessions), OCD compulsions score (CY-BOCS-compulsion), and standardised ADHD score (standardised ASRS or Conners scores) were used to predict change in YGTSS-TTSS at week 4.

### Video data analyses

As analysis of video data is extremely time consuming, we reduced the video data prior to statistical analysis as follows. For each week, 3 of a possible 5 daily recordings were selected at random for analysis. A video start time was randomly generated, and a 1min 50s video segment from commencing at this time point was selected for detail analysis. The same time points were assessed for each of the video sessions selected for that week. Videos collected during the 5 day baseline, and all videos collected from the waitlist group, were 5 minutes long and were processed as above. Videos collected during stimulation from the active and sham stimulation groups consisted of 5 minutes video recorded immediately prior to stimulation, 14 minutes of video recorded during while stimulation was delivered, and 5 minutes of video recorded after stimulation had ceased. For these videos, 1min 50s segments from each of these three time periods were randomly selected for assessment. Note, for clarification, in all cases in which a 1min 50s segment was selected from the 14 minute ‘stimulation’ period, this segment always contained 1min 50s during which stimulation was delivered, and commenced 10s after the start of the selected 2 minute period of stimulation.

This data reduction approach generated 5 minutes and 30 seconds of baseline video for each participant regardless of group. From the subsequent two-week period of recording, data reduction ideally resulted in 11 minutes of video for each participant in the waitlist group and 33 minutes for each participant in the sham and active groups. For each participant in the active and sham stimulation groups, the 33 minutes of video data would consist of 11 minutes prior to, 11 minutes during and 11 minutes after stimulation. The number of tics per minute (TPM) were calculated for each video recording session and the ‘*online’* effects of stimulation was assessed by calculating, for each session, the difference in TPM measured during stimulation from that measured immediately prior to stimulation. In this way, the online effects of stimulation were always referenced to current levels of tic frequency.

Tic counting (coding) was conducted by highly experienced researchers. The individuals who coded video recording sessions assessed 1.5 times the minimum number of participant videos per group x3 to create overlap between codes and permit assessment of coder reliability. Video assessment was performed by noting the time of every tic, together with information about what part of the body or type of vocalisation was involved. Importantly, coders were blind to the group membership of the participant, when the video was collected, and whether the video segment was from the period prior to stimulation, during stimulation, or post stimulation. Specifically, videos segments were provided to coders with a randomly generated name that did not contain any information about when the video was collected. The 1min 50s segments selected from each video session were randomised to create a video in which coders did not know whether the participant had already received stimulation, was receiving stimulation or had not yet received stimulation. Where possible, coders counted videos from participants they had been assessed during baseline, as they were familiar with that participant’s tics. Coder reliability was accepted at 80% agreement or greater, for each 1min 50s section of each video. If two coders had an agreement below 80% in any of the 1min 50s sections of a video, the two coders met and went through the video together until an agreement of +80% was reached. If +80% agreement was still not met, a third highly experienced researcher met with the coders and went through the video together until an agreement of +80% was reached. Once reliability had been achieved, an average of the tic counts was calculated for each section of each participant’s video. To increase reliability success, a highly experienced researcher met with the coders weekly to go through video sections in which movements and/or vocalisations were difficult to determine whether they were tics or voluntary actions.

### Ethics

This study was sponsored by the Nottingham University Hospitals NHS Trust and ethical approval was granted by the East Midlands -Leicester South Research Ethics Committee (22/EM/0024).

## Results

### Final sample characteristics

The final sample for statistical analysis comprised of a total 121 individuals with Tourette syndrome or Persistent Tic Disorder as follows: Active stimulation group (N=41); Sham stimulation group (N=39); or Treatment as usual (Waitlist) group (N=41). Tables 1 and 2 describe participant characteristics at baseline.

**Table 1:**
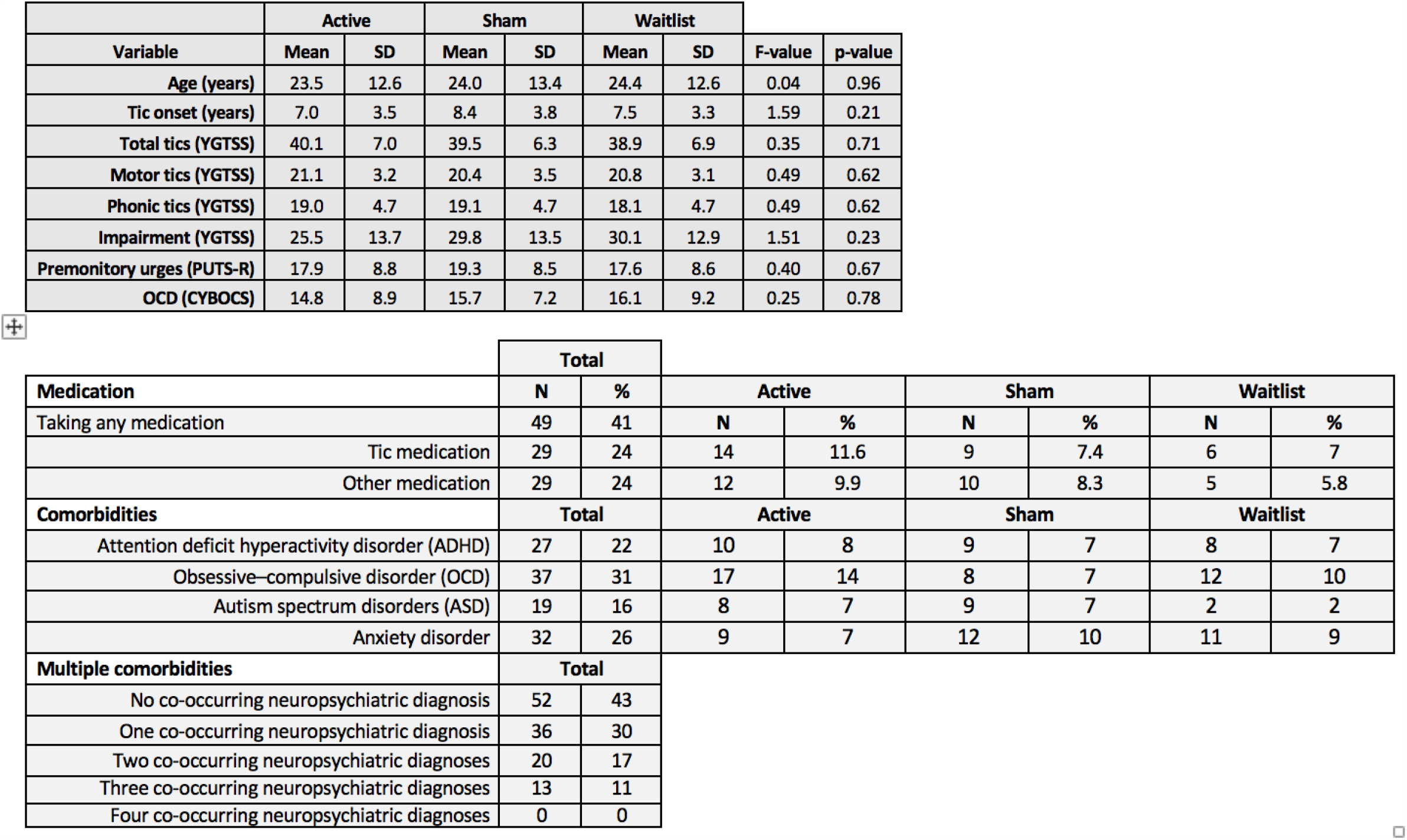
Upper panel. Values for age and primary outcome measures at baseline. Lower panel. Medication and co-morbidity details.

**Table 2:** Effect of 4-weeks of daily stimulation on key outcome measures for the active versus sham stimulation groups relative to a treatment-as-usual (no stimulation) control group.

| Contrast |  |  |  |  |  |
| --- | --- | --- | --- | --- | --- |
| Active vs. Sham |  |  |  |  |  |
| Outcomes | Observed difference | Effect-size | Low CI | High CI | p-value |
| YGTSS-TTSS | -5 | **-0.47** | -0.94 | -0.02 | **0.02** |
| YGTSS-motor | -2.03 | **-0.4** | -0.85 | 0.04 | **0.04** |
| YGTSS-phonetic | -3 | **-0.42** | -0.87 | 0 | **0.04** |
| YGTSS-impairment | 1.07 | 0.06 | -0.39 | 0.52 | 0.6 |
| PUTS-R | -0.59 | -0.05 | -0.5 | 0.39 | 0.4 |
| CYBOCS-total | -0.87 | -0.08 | -0.56 | 0.41 | 0.37 |
| Contrast |  |  |  |  |  |
| Active vs. Waitlist |  |  |  |  |  |
| Outcomes | Observed difference | Effect-size | Low CI | High CI | p-value |
| YGTSS-TTSS | -5 | **-0.48** | -0.97 | -0.04 | **0.02** |
| YGTSS-motor | -2.15 | **-0.43** | -0.88 | -0.01 | **0.03** |
| YGTSS-phonetic | -2.72 | **-0.42** | -0.85 | 0.02 | **0.04** |
| YGTSS-impairment | 1.86 | 0.11 | -0.35 | 0.58 | 0.7 |
| PUTS-R | -2.98 | -0.24 | -0.72 | 0.19 | 0.14 |
| CYBOCS-total | -3.17 | -0.25 | -0.75 | 0.22 | 0.15 |
| Contrast |  |  |  |  |  |
| Sham vs. Waitlist |  |  |  |  |  |
| Outcomes | Observed difference | Effect-size | Low CI | High CI | p-value |
| YGTSS-TTSS | -0.02 | 0.01 | -0.44 | 0.47 | 0.5 |
| YGTSS-motor | -0.13 | -0.03 | -0.47 | 0.42 | 0.44 |
| YGTSS-phonetic | 0.26 | 0.04 | -0.39 | 0.48 | 0.57 |
| YGTSS-impairment | 0.8 | 0.05 | -0.38 | 0.53 | 0.6 |
| PUTS-R | -2.39 | -0.19 | -0.65 | 0.26 | 0.2 |
| CYBOCS-total | -2.31 | -0.2 | -0.7 | 0.25 | 0.2 |

For each variable included, a one-way ANOVA was conducted to test for between-group differences at baseline. Inspection of Table 1 confirms that the groups did not differ in: age; number of years since tic onset; tic severity; or the severity of premonitory urge experiences. The final sample consisted of 62% males: Active group (26/41), Sham group (23/39), Waitlist group (26/41). A Chi-Square test confirmed that the numbers of males and females in each group did not differ significantly (X^2^ = 0.024, df=2, p = 1.0).

### Efficacy of sham stimulation

Prior to commencing the trial, we had piloted the sham stimulation condition to confirm that it differed from active stimulation. Specifically, we used MEG (magnetoencephalography) to investigate whether sham stimulation led to the entrainment of mu-band brain oscillations that we observe during active stimulation (Houlgreave et al., 2022). We were able to confirm that, in contrast to active stimulation, mu-band sham stimulation delivered at 50% of threshold produces no change in mu-band power or phase-synchrony.

At the conclusion of the trial, participants in both the active and sham stimulation groups were asked if they knew which condition they had been assigned to. All confirmed that they did not. They were also asked if they could tell which group they had been assigned to. The majority reported that they had no idea which condition they had been assigned to, but some reported that they had guessed which condition they took part in. To determine if these participants were correctly able to identify in which condition they had taken part, we conducted a Fisher’s Exact Test. Overall, there was a bias for those participants who guessed which condition they were in to report that they had taken part in the sham stimulation group. 64% of those who guessed and were assigned to the active stimulation group, incorrectly guessed they were in the sham group. By contrast, 40% of those who guessed and were assigned to the sham stimulation group, incorrectly guessed they were in the active group. The Fisher’s Exact Test revealed an Odds Ratio of 0.38, [CI: 0.1 - 1.3], p = 0.22, which indicated that there was no evidence that to indicate that participants could reliably identify which type of stimulation they received.

### Medication and co-occurring neuropsychiatric conditions

Details of participants’ medication and co-occurring neuropsychiatric diagnoses are presented in Table 1. The number of participants who presented with a Tourette syndrome of Chronic tic disorder diagnosis but with no co-occurring neuropsychiatric conditions was 52 (43%). The remaining 57% of participants presented with between one and three co-occurring diagnosed neuropsychiatric conditions. The most common co-occurring neuropsychiatric conditions were: Obsessive-compulsive disorder (OCD) [31%]; anxiety disorder [26%]; attention deficit hyperactivity disorder (ADHD) [22%]; and, autism spectrum disorder (ASD) [16%]. A small number of those with anxiety disorder also presented with other neuropsychiatric conditions that included: depression and functional neurological disorder. The number of participants with co-occurring conditions was evenly distributed across the three conditions (see Table 1).

A number of participants 49/121 (41%) were taking prescribed medication for a neuropsychiatric condition during the trial. 29/121 (24%) were taking a recognised tic medication, whereas 29/121 (24%) were taking a medication prescribed for another neuropsychiatric condition (e.g., for ADHD, anxiety, or depression). Full details of medication are available on request.

### Primary outcomes: Change in tic severity, premonitory urges, and OCD symptoms after 4 weeks of daily stimulation

Group means and standard deviations for YGTSS-motor, YGTSS-phonic, YGTSS-impairment, PUTS-R, and CYBOCS scores at baseline are presented in Table 1. The baseline mean YGTSS-TTSS score for the <u>entire sample</u> was 39.0 ± 6.7 (SD), and the median score was 40.0 ± 9 (IQR). The 95% confidence interval ranged from 26 to 48 and the 25^th^ percentile value was 26. Based upon these data we estimated that a reduction in 4 points represented a 25% decrease in tic severity based upon YGTSS-TTSS. Identical analyses were also conducted for YGTSS-motor, YGTSS-phonic, YGTSS-impairment, PUTS-R, and CYBOCS scores to estimate the magnitude of a 25% reduction in each score.

Participants were removed from further analysis if they exhibited a YGTSS-TTSS at baseline (week 0) below the inclusion threshold (i.e., a YGTSS-TTSS of 15 or lower) or if there was missing data for week 4. This led to the removal of 4 participants in total, leaving 39 participants in each group. Figure 2A illustrates the changes in total tic severity (YGTSS-TTSS) observed at week 4, relative to baseline levels, for each group. Inspection of this figure indicates a reduction in mean value by week 4 for the Active condition. By the end of the 4-week stimulation phase of the trial, the Active stimulation group exhibited a mean reduction in YGTSS-TTSS of 7.13 (1.1 SD units) points compared to a mean reduction of 2.13 (0.32 SD units) points for the Sham stimulation group and 2.26 (0.34 SD units) points for the Waitlist (no stimulation) groups.

**Figure 2:**
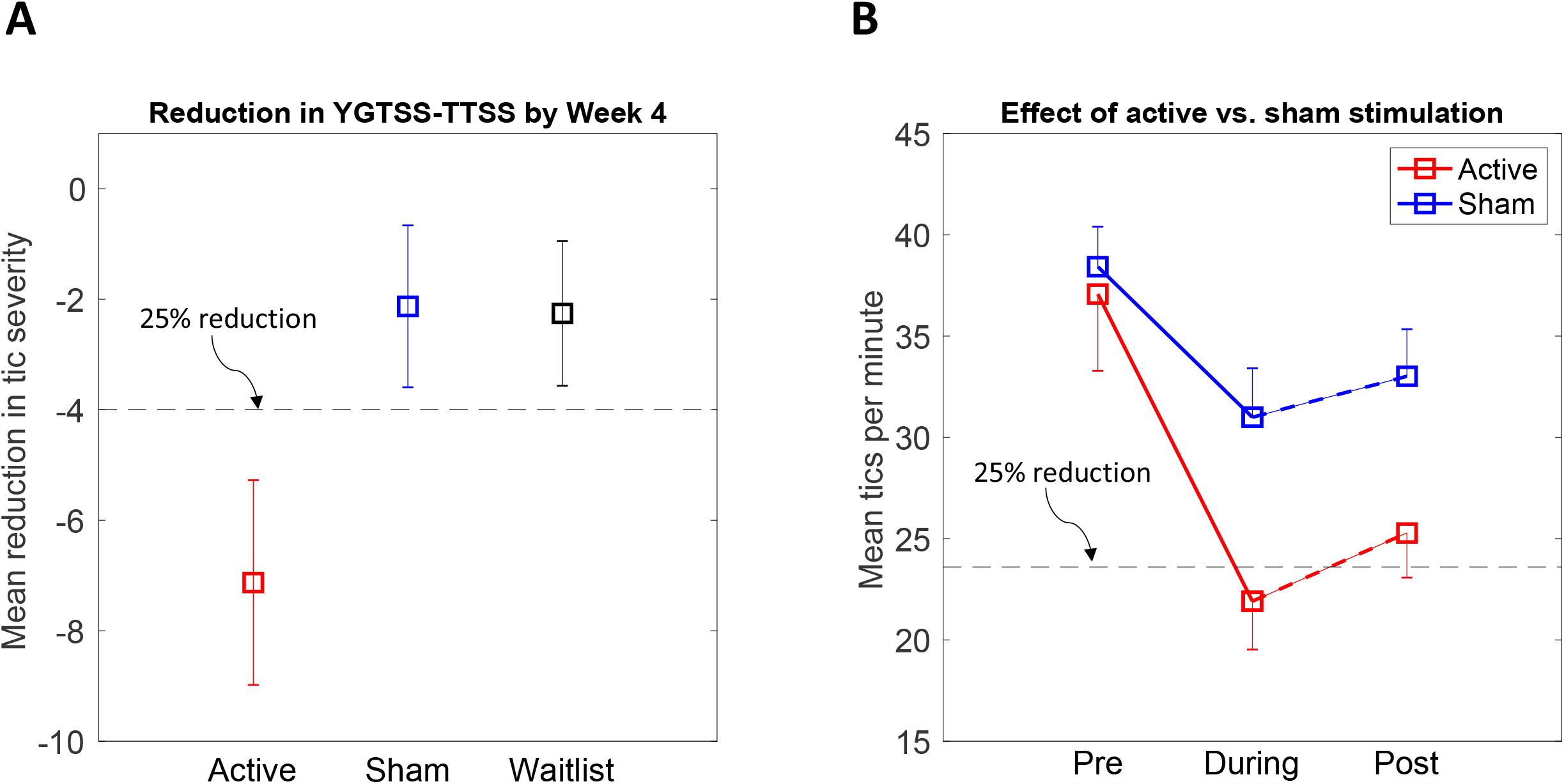
**A**. Mean reduction in total tic severity score (YGTSS-TTSS) for each group after 4 weeks of stimulation relative to baseline levels. The dotted line represents a 25% reduction in YGTSS-TTSS. Error bars are standard errors. **B**. Mean reduction in tics per minute (TPM) for active and sham stimulation sessions immediately prior to, during, and post stimulation. The dotted line represents a 25% reduction in TPM. Error bars are standard errors.

Statistical testing of the between-group differences in the magnitude of the change from baseline scores at week 4, calculated separately for each individual, was performed using *a priori* planned non-parametric permutation testing. These analyses revealed that: (a) the magnitude of the <u>reduction</u> in YGTSS-TTSS for the Active stimulation group was significantly greater than that observed for the Waitlist control group (observed difference = -4.9, Effect size = -0.5, p = 0.02); (b) the mean YGTSS-total score for the Sham stimulation group did <u>not</u> differ from that of the Waitlist control group (observed difference = 0.13 Effect size = -0.02, p = 0.5), and, most importantly, (c) the difference in means for the Active and Sham groups was substantial and significantly different (observed difference = -5, Effect size = -0.48, p < 0.02). A similar pattern was also observed for YGTSS-motor and YGTSS-phonic scores separately, but was <u>not</u> observed for YGTSS-impairment, PUTS-R, or CYBOCS-total scores. Relevant results are summarised in Table 2. Additional analyses using mixed 3×5 ANOVAs confirmed that for all of the assessment measures, there was a statistically significant reduction in symptom severity across the 4-week period (minimum F[4,456] = 2.9 [range 2.9 – 10.4], p < 0.025).

### Differences in the number of Individuals in each group who exhibited a clinically meaningful response

Based upon previously published studies (e.g., Jeon et al., 2013) we determined that the criterion for a positive, clinically meaningful, reduction in tic severity in response to stimulation would be a reduction of 25% or greater in the primary outcome measure, YGTSS-TTSS, by the final week of the 4-week stimulation period. A similar criterion of a 25% reduction in clinical score was also adopted for other the outcome measures. Table 3 presents group differences (i.e., Active vs. Sham stimulation) in the number and percentage of responders identified when applying the 25% response criteria to YGTSS-TTSS scores. Analyses revealed that 59% of the Active stimulation group were responders by these criteria, compared to 33% of the Sham stimulation group. Statistical testing of this contrast revealed that the number of responders in the Active stimulation group was substantially greater than the number observed in the Sham stimulation group (Odds ratio = 2.9, CI = 1.1 - 7.2). This analysis also confirmed that there was a positive association (Cramer’s V = 0.2) between receiving Active stimulation and exhibiting a clinically meaningful reduction (25%) in tic severity, compared to Sham stimulation, and the relative likelihood of Active stimulation leading to a meaningful positive was 67%. The number of responders observed in the Active stimulation versus Sham stimulation groups, and key statistics, for other outcome measures are reported in Table 3.

**Table 3:** Shows the number of responders (i.e., individuals who exhibit a clinically meaningful reduction) for Active vs. Sham stimulation for each primary outcome measure. We also report the results of the statistical analyses (Odds ratio - Fisher’s Exact Test). CI = 95% confidence intervals. RRR = Relative risk reduction which indicates relative likelihood of being a responder given Active vs. Sham stimulation.

|  | Responders |  | Non-responders |  |  |  |  |  |
| --- | --- | --- | --- | --- | --- | --- | --- | --- |
| **YGSS-TTSS** | **N /39** | **%** | **N /39** | **%** | **Odds ratio** | **low CI** | **high CI** | **RRR (%)** |
| Active | 23 | 59.0 | 16 | 41.0 | **2.9** | 1.1 | 7.2 | **67** |
| Sham | 13 | 33.3 | 26 | 66.7 |  |  |  |  |
| **YGSS-motor** | **N /39** | **%** | **N /39** | **%** | **Odds ratio** | **low CI** | **high CI** | **RRR (%)** |
| Active | 30 | 76.9 | 9 | 23.1 | **2.6** | 1 | 6.85 | **67** |
| Sham | 22 | 56.4 | 17 | 43.6 |  |  |  |  |
| **YGSS-phonetic** | **N /39** | **%** | **N /39** | **%** | **Odds ratio** | **low CI** | **high CI** | **RRR (%)** |
| Active | 21 | 53.8 | 18 | 46.2 | **2.3** | 0.9 | 5.8 | **51** |
| Sham | 13 | 33.3 | 26 | 66.7 |  |  |  |  |
| **YGSS-impairment** | **N /39** | **%** | **N /39** | **%** | **Odds ratio** | **low CI** | **high CI** | **RRR (%)** |
| Active | 13 | 33.3 | 24 | 61.5 | 0.9 | 0.4 | 2.3 | 5.7 |
| Sham | 14 | 35.9 | 23 | 59.0 |  |  |  |  |
| **PUTS-R** | **N /39** | **%** | **N /39** | **%** | **Odds ratio** | **low CI** | **high CI** | **RRR (%)** |
| Active | 21 | 53.8 | 19 | 48.7 | 1.1 | 0.5 | 2.7 | 5 |
| Sham | 19 | 48.7 | 19 | 48.7 |  |  |  |  |
| **CYBOCS-total** | **N /32** | **%** | **N /36** | **%** | **Odds ratio** | **low CI** | **high CI** | **RRR (%)** |
| Active | 19 | 59.4 | 13 | 40.6 | 0.8 | 0.6 | 4.1 | 10 |
| Sham | 23 | 63.9 | 13 | 36.1 |  |  |  |  |

### Moderation of change in tic severity outcome by co-occurring conditions

Tic disorders such as Tourette syndrome and Chronic tic disorder are invariably accompanied by one or more co-occurring neuropsychiatric conditions, most often OCD, ADHD, and anxiety disorder. This was certainly the case for the individuals participating in this study (see Table 1) where only 43% of the sample presented without a co-occurring neuropsychiatric diagnosis. The remaining 57% presented primarily with co-occurring OCD (31%), anxiety disorder (26%) and ADHD (22%), and it is possible that symptom severity for one or more of these conditions moderated the effects of rMNS, and influenced the magnitude of tic reduction observed after 4 weeks of stimulation. To investigate this we conducted a stepwise multiple regression analysis in which the following measurements, collected at baseline (week 0), were used to predict YGTSS-TTSS at week 4: Study condition; CYBOCS-total; CYBOCS-obsessions; CYBOCS-compulsions; standardized ADHD score; Beck Anxiety Inventory (BAI) score. This analysis revealed that the factors Study condition (coefficient = 2.7, t-value = 2.5, p-value < 0.02) and CYBOCS-total (coefficient = -0.2, t-value = -2.1, p-value < 0.05) were both significant predictors of YGTSS-TTSS and week 4 (F = 5.0, R2 = 0.08, p < 0.01). No other outcome measures (e.g., standardised ADHD score or BAI score) contributed to the regression model.

### Online effects of rMNS on tic frequency

The results for the tic severity outcome measures reported above provide positive evidence for a clinically meaningful ‘offline’ or treatment effect of rMNS (i.e., a reduction in tic severity that outlasts any period of stimulation). However, in response to individuals stating that what they really wanted was increased control over their tics, specifically in situations where they felt it important not to tic, a key objective of our research was to develop a wearable non-invasive stimulation approach that could be used outside of the clinic and which produced an *immediate* reduction in tics (i.e. tic frequency) whenever stimulation was applied, i.e., an ‘online’ rather than ‘offline’ effect of stimulation. To investigate this effect we assessed tic frequency (i.e., number of tics per minute [TPM]) during video recording sessions during which Active stimulation was delivered compared with video recording sessions in which Sham stimulation was delivered. For each session, the change in tic frequency was calculated by subtracting TPM in the period during stimulation with TPM measured during the 5 minute period immediately prior to stimulation.

Current video data analyses are based on 82 video sessions from 16 participants receiving active stimulation and 91 video sessions from 17 participants receiving sham stimulation.

### Group characteristics

The group undertaking the video analyses were a random sub-group of the larger trial group. The characteristics of those participants included in the video analyses (i.e., all those analysed to date) are shown in Table 4. Those in the Active and Sham stimulation groups do not differ in age, age at tic onset, tic severity, or severity of premonitory urges.

**Table 4.**
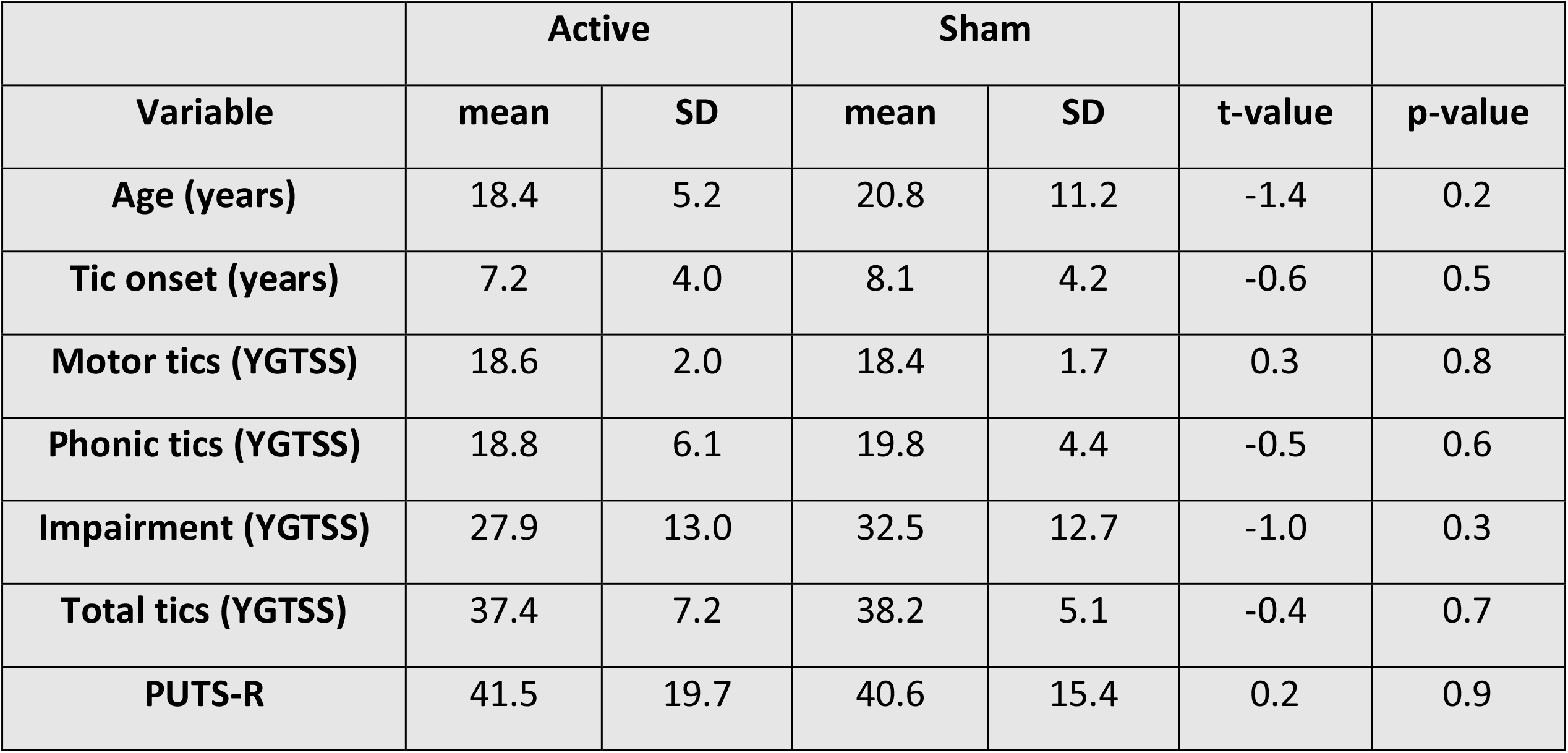
Characteristics of individuals who provided video sessions for analysis.

#### Baseline tic frequency characteristics

Tic severity for the entire group was assessed based upon video recordings collected in the week prior to stimulation commencing (Week 0). These data confirmed that the group exhibited a large number of tics per minute (TPM) [mean = 44.1 ± 29.1 TPM; median = 39.3 IQR = 32.1]. The 5^th^ percentile value was 11.9 TPM and the 1st percentile value was 6.3 TPM. Based upon these data we set a tic frequency minimum cut-off of 6 TPM (i.e., for a video session to be included in the analyses the individual must exhibit a tic frequency of 6 TPM or greater in the period immediately prior to stimulation.

We again adopted a 25% reduction in tic frequency as a clinically meaningful positive treatment response and used this value as our criterion for defining those individuals who exhibit a positive response to stimulation. Based upon the baseline data from our sample, a 25% reduction in tic frequency would equate to a reduction of 15.3 TPM.

### Effects of active vs. sham stimulation on tic frequency

The mean number of TPM for Active versus Sham stimulation sessions at each period are presented in Figure 2B. Inspection of this figure suggests that the stimulation groups were equivalent in the period immediately prior to stimulation being delivered, but differed during stimulation and post stimulation. To investigate this we conducted a mixed 2×3 ANOVA with the between-subject factor of Stimulation type (Active vs. Sham) and the within-subject factor of Period (pre-stimulation, during stimulation, post-stimulation). The ANOVA revealed significant main effects of Stimulation type (F[1,177] = 4.0, p < 0.05) and Period (F[2,354] = 23.0, p < 0.0001). The Stimulation type x Period interaction was marginally significant (F[2,354] = 2.8, p = 0.06).

To further explore group differences, we conducted a set of planned contrasts using non-parametric permutation tests to investigate the effect of Stimulation type at each period. These contrasts revealed that while the Stimulation type groups did not differ immediately prior to stimulation being delivered (observed difference = -1.4, effect size = -0.05, p-value = 0.8), the Active stimulation group exhibited significantly fewer TPM than the Sham stimulation group both during the stimulation period (observed difference = -9.1, effect size = -0.4, p-value < 0.01) and post stimulation (observed difference = -7.7, effect size = -0.36, p-value < 0.02).

### Analyses of difference measures

For completeness, we also calculated for each individual, the difference in tic frequency (i.e., TPM) during stimulation compared to that individual’s pre-stimulation tic frequency. The mean reduction in tic frequency for the Active group was -15.6 ± 36.6 TPM and for the Sham group was -7.7 ± 17.3 TPM. This difference represents a small-medium sized effect (Effect size [standardised mean difference] = - 0.3, CI =-0.02 to 0.58. Non-parametric permutation testing also confirmed that this difference was statistically significant (p = 0.03). Note that the mean reduction for the Active stimulation group exceeded the response criterion for a ‘clinically meaningful’ reduction in tic severity (i.e., a 25% reduction in tics equalling -15.3 TPM) while the Sham stimulation group did not.

## Discussion

The starting point for this study was the recognition that TS and tic disorders represent an area of considerable unmet clinical need - with many individuals finding it very difficult to access effective treatments - and that patients are looking for a safe and effective non-drug treatment for controlling their tics, and suppressing the urge-to-tic, that can be used outside of the clinic within a community setting. In particular, individuals with TS often state that they want a treatment that gives them immediate <u>control over their tics, on demand</u>, and thus provides them with more choice over when and where they might express their tics.

Morera Maiquez and colleagues had previously demonstrated in a small open-label study that brief trains of rhythmic mu-band median nerve stimulation (MNS) delivered to the right wrist was effective, relative to no stimulation, in reducing both tic frequency and tic intensity in individuals with TS (Morera Maiquez et al., 2020). We now conducted a larger, UK-wide, double-blind, sham-controlled, study of the efficacy of home-administered rhythmic mu-band MNS.

The results of this study can be summarised as follows. First, blind analyses of video recordings of stimulation sessions demonstrated that <u>tic frequency</u> (tics per minute) reduced substantially during Active stimulation. Importantly, this reduction was significantly larger than that observed during Sham stimulation. Furthermore, the ‘online’ reduction in tic frequency that was observed during active stimulation sessions represents a clinically meaningful reduction in tic frequency (i.e., a mean reduction in tic frequency of at least 25%). Second, four weeks of daily sessions of 10-minutes of active MNS stimulation led to a substantial, clinically meaningful (i.e., an average reduction of at least 25%), and statistically significant ‘offline’ reduction in <u>tic severity</u>, as measured by the YGTSS-TTSS measure. Specifically, total tic severity reduced by an average of 7.13 points or 1.06 standard deviation units for the Active stimulation group relative to baseline (this amounts to a 35% change based upon whole sample baseline measurements). By contrast, average tic severity only reduced by 2.13/2.11 points or 0.32/0.31 standard deviation units for the Sham stimulation and Treatment as usual control groups. Third, the difference in the magnitude of the reduction in tic severity (YGTSS-TTSS) for the Active stimulation group was substantially larger, clinically meaningful (effect size = 0.5), and statistically significant (p = 0.02) compared to both the Sham stimulation and Treatment as usual control groups, which did not differ from one another. Finally, a substantial majority (59%) of participants receiving 4 weeks of active stimulation exhibited a ‘*clinically meaningful reduction in tic severity’*, and statistical testing confirmed that the relative likelihood (Relative Risk Ratio) of Active stimulation leading to a ‘*clinically meaningful reduction in tic severity’* was 67%. These findings are discussed below.

### ‘Online’ effects of rhythmic median nerve stimulation on tic frequency

As noted above, Morera Maiquez and colleagues had previously demonstrated in an open-label study that rhythmic trains of mu-band median nerve stimulation (MNS), delivered at the wrist, were effective in reducing tic frequency and tic intensity in individuals with TS, relative to a no stimulation control condition (Morera Maiquez et al., 2020). That study demonstrated that rhythmic mu-band MNS was sufficient to produce an overall reduction of 31% in tic frequency and a 30% reduction in the self-estimated urge-to-tic. Support for this finding was recently obtained in an independent study that examined the efficacy of rhythmic MNS in reducing tic frequency, tic intensity and self-estimated urge-to-tic ratings in comparison to an arrhythmic MNS control condition (Iverson, Arbuckle, Ueda, Song, Bihun, Koller, et al., 2023). Importantly, this study replicated the findings of the Morera Maiquez study and confirmed that tic frequency and the urge-to-tic were both significantly reduced during rhythmic MNS, in some cases producing a dramatic reduction. However, as the authors point out, without an appropriate sham control condition, it is not possible to exclude a placebo effect. In the current study we directly compared rhythmic mu-band MNS against a sham control condition (i.e., identical stimulation but delivered at 50% of threshold). Our results demonstrate for the first time in a sham-controlled study that rhythmic mu-band MNS is sufficient to substantially reduce tic frequency in a group of individuals with TS or chronic tic disorder, and that the magnitude of this reduction in tic frequency is clinically meaningful and significantly greater than that observed for sham stimulation.

Iverson et al. directly compared the effects of rhythmic mu-band MNS with a matched arrhythmic MNS control condition, in which an identical number of MNS pulses were delivered but in an arrhythmic rather than a rhythmic pattern. They reported that the beneficial effects of MNS on tic frequency were equivalent, insofar as both rhythmic and arrhythmic MNS substantially reduced tic frequency, intensity and the urge-to-tic by a similar amount, relative to a no stimulation control condition. This is an interesting finding given that previous studies have compared rhythmic versus arrhythmic MNS using a number of approaches and found them to have different effects. First, using both EEG and MEG recording methods, it was shown that pulse trains of rhythmic MNS are effective in entraining sensorimotor mu-band oscillations (i.e., increasing the amplitude and phase synchronisation of oscillations at the frequency of stimulation), whereas pulse trains of arrhythmic MNS do not (Houlgreave, Morera Maiquez, Brookes, Jackson, 2022; Morera Maiquez et al., 2020). Second, in two separate studies that compared the effects of rhythmic versus arrhythmic MNS on the initiation of volitional movements it was demonstrated, in both studies, that rhythmic MNS slowed movement initiation compared to arrhythmic MNS (Morera Maiquez et al., 2020). Nonetheless, the finding by Iverson et al. that stimulation using rhythmic and arrhythmic mu-band MNS produces comparable decreases in tic frequency indicates that, while entrainment of mu-band oscillations during rhythmic mu-band MNS may be *sufficient* to produce a substantial reduction in tic frequency in TS, it may not a *necessary* condition for reducing tics using MNS. It is clear that further investigation will be necessary to better understand how MNS can be optimised to produce the most effective reduction in tic frequency, intensity and the urge-to-tic. One additional finding reported in the Iverson et al. study was that they reported that the benefits of MNS did not persist beyond the period of stimulation. We address this issue below when we consider the beneficial ‘offline’ effects of repeated daily sessions of MNS over a 4-week period.

### ‘Offline’ treatment effects of daily rhythmic median nerve stimulation on tic severity

While the starting point of this study was to investigate the clinical benefits of home-administered ‘online’ MNS in comparison to a sham-MNS control condition, it is obvious that an effective ‘offline’ treatment effect of MNS (i.e., a clinical benefit of MNS that persists after stimulation has ceased) would be an extremely positive outcome. In the current study, participants received five daily 14-minute sessions (each containing 10 minutes of stimulation) of median nerve stimulation each week over a period of four weeks, and clinical outcome measures were obtained weekly, with tic severity (YGTSS-TTSS) being the primary outcome measure. The results of the study demonstrated that after 4 weeks of stimulation there was a substantial, clinically meaningful (i.e., an average reduction of at least 25%, Jeon et al., 2013), and statistically significant reduction in total tic severity (YGTSS-TTSS) of 7.1 points which was not observed for a control group who received sham-stimulation or an additional ‘waitlist’ control group who received treatment as usual. Importantly, the difference in the magnitude of the reduction in total tic severity the active stimulation group was substantially larger, and statistically significant (p = 0.02), compared to both the sham stimulation and waitlist control groups.

It is worth noting that the magnitude of the reduction in tic severity (i.e., a standardised mean difference of 0.5) observed in the current study compares favourably with recent studies evaluating the efficacy of behavioural interventions for tic disorder. For example, a recent study by Hollis and colleagues reported a multi-centre, parallel-group, single-blind, randomised controlled trial (ORBIT) that compared 10-weeks of therapist-supported online remote behavioural treatment (i.e., Exposure and Response Prevention – ERP) against a psychoeducation control group (Hollis, Hall, Jones, Marston, Le Novere, Hunter, et al., 2021). This study demonstrated a reduction in total tic severity score of 4.5 points following ERP compared to a reduction of 1.6 points for the psychoeducation group, and a standardised mean difference of 0.31. Similarly, a study reporting a trial of a different behavioural intervention - Comprehensive Behavioural Intervention for Tics (CBIT) - compared against a psychoeducation control group, demonstrated a reduction in total tic severity score of 7.6 points following 10-weeks of CBIT compared to a reduction of 3.5 points for the Psychoeducation group and a standardised mean difference of 0.68 (Piacentini, et al., 2010). Finally, a study reporting a trial of 8 sessions (10-weeks) of CBIT compared against a group receiving 8 sessions of supportive treatment demonstrated a reduction of 6.2 points following CBIT compared to a reduction in total tic severity score of 2.5 points following supportive treatment, and a standardised mean difference between the groups of 0.57 (Wilhelm, Peterson, Piacentini, Woods, Deckersbach, Sukhodolsky, et al., 2012).

### Limitations of the study

The current study has a number of limitations that might be addressed in further investigations. First, as it is the first double-blind sham-controlled trial of home-administered MNS for tic disorders it would be prudent to replicate the effects reported here. Second, the current study was designed in response to the desire, often stated by patients, for a means of controlling their tic symptoms on demand, and was optimised to extend our previous investigations of the ‘online’ effects of rhythmic mu-band MNS. For this reason, the daily periods of active or sham stimulation were relatively brief in order to facilitate the repeated collection of video recordings. While we did obtain a clear treatment effect of active stimulation compared to sham stimulation on tic severity, we feel that it is likely that increasing the daily duration of active stimulation (e.g., from 10 minutes to 30 minutes or 45 minutes) would have led to an even larger reduction in tic severity. We are currently conducting follow-up studies in which we are investigating the benefits of longer durations of stimulation of tic severity. Third, blind analysis of tics from video recordings is complex and very time-consuming, as analysis of each minute of video can take more than one hour to complete. In the current study, we opted for a data reduction strategy in which we randomly sampled video segments for blind analysis in which all of the tics occurring within that segment were counted by more than one expert assessor. Despite implementing this data reduction strategy, the tic-counting analyses reported in this study took four individuals over 6 months to complete. One limitation of simply counting the number of tics per minute that are observed is that this measure does not adequately capture any qualitative changes in tic severity or complexity that may occur in response to treatment. For this reason, further investigation of likely qualitative changes in tic severity and complexity that arise during active stimulation should be pursued. Finally, the study involved home-administration of a wearable stimulation device that was positioned over the median nerve of the right wrist during use. In particular, it is possible that the individual threshold for effective stimulation altered for some participants over the course of the 4-week stimulation period. One consequence of this could be to reduce the effectiveness of the pre-programmed level of stimulation. Participants had received comprehensive training in how to wear the device and their individual stimulation threshold was programmed into the device prior to them taking the device home and commencing the 4-week stimulation period.

Nonetheless, unlike in a laboratory or clinical setting, we had minimal control over these factors once the participant was using the device in their home environment.

## Conclusion

We conducted a UK-wide, parallel, double-blind, sham-controlled evaluation of the efficacy of home-administered rhythmic median nerve stimulation as a potential treatment for tic disorders. Importantly, stimulation was delivered at the wrist my means of a wearable and unobtrusive ‘watch-like’ stimulation device. The primary outcome measure for ‘offline’ (i.e., treatment) effects of stimulation was tic severity (YGTSS-TTSS). The primary outcome measure for ‘online’ (i.e., during stimulation) effects of stimulation and was a reduction in tic frequency during stimulation.

The study has demonstrated that 10 minutes of active rhythmic mu-band MNS, delivered daily over a 4-week period, was sufficient to substantially reduce tic severity in individuals with tic disorders, and that tic frequency reduced substantially while stimulation was delivered. Importantly, the reduction in both tic severity and tic frequency observed for active stimulation differed substantially from that observed for sham stimulation. It should be noted that the stimulation reported in this study is suitable for use with children and young adults, can be delivered by means of a wearable ‘watch-like’ device worn on the wrist, that can readily be used outside of the home, for example in a school or workplace setting.

## Data Availability

The data that support the findings of this study will be available on request from the corresponding author [SRJ]. The data are not publicly available due to ethical restrictions as they contain information that could compromise the privacy of research participants.

## Acknowledgements

This work was supported by Tourettes Action (UK), The Tourettes Association of America, The Medical Research Council, and the NIHR Nottingham Biomedical Research Centre. The views expressed are those of the authors and not necessarily those of the NHS, the NIHR or the Department of Health. We thank Tourettes Action (UK) for assisting with participant recruitment and Chris Hollis, Eileen Joyce, and Jeremy Stern for providing clinical guidance on prescribed medications for tic disorders, and we thank Neurotherapeutics Ltd for providing the Neupulse devices used in this trial.

